# Racial discrimination, low trust in the health system, and COVID-19 vaccine uptake: a longitudinal observational study of 633 UK adults from ethnic minority groups

**DOI:** 10.1101/2021.08.26.21262655

**Authors:** Elise Paul, Daisy Fancourt, Mohammad Razai

## Abstract

**Objective:** To examine whether racial/ethnic discrimination predicts future COVID-19 vaccine refusal, and whether this association is explained by trust in government and the health system.

**Design:** Longitudinal observational study of racial/ethnic discrimination occurring since the start of the first lockdown (measured in July 2020) and later COVID-19 vaccine status.

**Setting:** UK (England, Scotland, Wales, and Northern Ireland)

**Participants:** 633 adults belonging to ethnic minority groups who took part in the UCL COVID-19 Social Study.

**Main outcome measure:** COVID-19 vaccine refusal (vs accepted/waiting/had at least one dose) between 23 December 2020 and 14 June 2021.

**Results:** Nearly one in ten (6.7%) who had refused a COVID-19 vaccine had experienced racial/ethnic discrimination in a medical setting since the start of the pandemic and had experienced twice as many incidents of racial/ethnic discrimination than those who had accepted the vaccine. Structural equation modelling results indicated a nearly 4-fold (odds ratio [OR] = 3.9, 95% confidence interval [CI] = 1.4 to 10.9) total effect of racial/ethnic discrimination on refusing the vaccine was which was mediated by low trust in the health system to handle the pandemic (OR = 2.5, 95% CI = 1.1 to 5.4). Analyses adjusted for a range of demographic and COVID-19 related factors.

**Conclusions:** Findings underscore the importance of addressing racial/ethnic discrimination and the role the National Health Service in regaining trust from ethnic minority groups to increase COVID-19 vaccine uptake amongst ethnic minority adults.

## Introduction

Despite the relative overall success of the UK’s vaccination programme, the differential uptake of COVID-19 vaccines is a cause for concern. The most recent data from the Opinions and Lifestyle Survey showed that as of 18 July 2021, vaccine uptake in eligible adults was lower in ethnic minority groups as a whole (80%) compared to White British (91%), particularly in people self-identifying as Black or Black British (68%), and in people of mixed ethnicity (79%).^1^ Additionally, data show higher COVID-19 vaccine hesitancy (a delay in acceptance or refusal of safe vaccines despite availability of vaccine services)^2^ in ethnic minority groups (9%), particularly in Black or Black British adults (21%) compared to White British adults (4%).^1^ Further, large UK surveys of attitudes towards COVID-19 vaccination have shown much greater proportions of vaccine hesitancy in some of these ethnic minority groups.^3–5^ This is concerning as ethnic minorities have been disproportionately and severely impacted by COVID-19 with higher rates of infection, hospitalisation and death rates compared to White ethnicities.^6,7^ Further, a refusal rate of more than 10% could also undermine control of the current pandemic and achieving population level immunity.^8^

Though the reasons for vaccine hesitancy amongst ethnic minority groups are complex and multifactorial, racial discrimination is likely to be a key upstream cause.^9,10^ Research conducted before the COVID-19 pandemic has found that racial discrimination contributed to delayed medical screenings,^11^ lower expectations of the quality of medical treatment,^12^ and barriers to seeking mental and physical health care services.^13^ In relation to COVID-19 vaccination, a cross-sectional study in the USA conducted in December 2020, found lifetime experiences of racial discrimination – but not discrimination due to religion, gender, or sexual orientation – was associated with 21% increased odds of COVID-19 vaccine hesitancy.^14^ Other survey data confirms the influence of racial and ethnic discrimination with COVID-19 vaccine hesitancy.^15,16^

Racial and ethnic discrimination in turn leads to mistrust of government and public health institutions,^17,18^ which are key barriers to vaccination.^9,10,16,19^ Qualitative research conducted with ethnic minority groups during the third UK lockdown also points to mistrust of government and public health bodies as a mediator between racial/ethnic discrimination and vaccine hesitancy.^20^ Further, quantitative studies have suggested that once trust in government and the health system are accounted for, ethnic minority group status no longer associates with COVID-19 vaccine unwillingness.^21,22^ Longitudinal and cross-sectional studies have highlighted that the main reasons for COVID-19 hesitancy among ethnic minorities centre around trust, with Black ethnicities substantially more likely to state that they “don’t trust vaccines” compared with White people (29.2% vs 5.7%).^4^ Lack of trust can be self-perpetuating as it can lead to lower participation in research amongst ethnic minority groups, in particular COVID-19 research. This may in turn lead to a paucity of data on COVID-19 vaccines amongst ethnic minorities and exacerbate low confidence in the vaccines amongst these groups.^23^

To address differential uptake of COVID-19 vaccines amongst ethnic groups in the UK, public health messaging aimed at ethnic minorities emphasised the importance, necessity and safety of vaccines.^24^ Additionally, places of worship were used as ‘pop up’ vaccination sites and high-profile ethnic minority celebrities issued an open letter to raise confidence in COVID-19 vaccines.^25,26^ However, these efforts have been insufficient to prevent inequalities in vaccine uptake. Therefore, more work is urgently needed to mitigate the unequal and severe effects of the pandemic on ethnic minority populations. The objective of this study is therefore to examine the longitudinal associations between experiences of racial/ethnic discrimination and COVID-19 vaccine refusal and explore whether low trust in government and the health system could explain this association. This work could help to guide future interventions to support vaccine uptake amongst ethnic minority groups.

## Methods

### Participants

We used data from the COVID-19 Social Study; a large ongoing panel study of the psychological and social experiences of over 70,000 adults (aged 18+) in the UK during the COVID-19 pandemic. The study commenced on 21 March 2020 and involves online weekly (to August 2020) then monthly (four-weekly) data collection for the duration of the pandemic. Sampling is not random and therefore is not representative of the UK population, but the study does contain a heterogeneous sample. The sample was recruited using three primary approaches. First, convenience sampling was used, including promoting the study through existing networks and mailing lists (such as large databases of adults who had previously consented to be involved in health research across the UK), print, and digital media coverage. Second, more targeted recruitment was undertaken focusing on (i) individuals from a low-income background, (ii) individuals with no or few educational qualifications, and (iii) individuals who were unemployed. Third, the study was promoted via partnerships with third sector organisations to vulnerable groups, including adults with pre-existing mental health conditions, older adults, carers, and people experiencing domestic violence or abuse.

Participants who took part in the study between 23 July 2020 to 14 June 2021 (n = 46,991) were eligible for inclusion. We excluded participants if they had missing data on any study variables. A total of 36,883 had non-missing vaccination status data collected from 23 December 2020 onwards. Of these, 22,212 also had non-missing data on the discrimination module, which was administered the week of 23 to 30 July 2020, and 21,636 also had non-missing data on all other study variables. Of these, 712 responded at the baseline interview that they belonged to an ethnic minority group (Asian/Asian British - Indian, Pakistani, Bangladeshi, other; Black/Black British - Caribbean, Africa; Mixed race - White and Black/Black British; mixed race-other; Chinese/Chinese British; Middle Eastern/Middle Eastern British – Arab, Turkish, other; or other ethnic group]). As our focus was on COVID-19 vaccine refusal versus acceptance, we eliminated the 79 ethnic minority group adults who met all other inclusion criteria but who had not yet been offered the vaccine, leaving a final sample of 633. See Supplemental Table S1 for a comparison of excluded and included participants.

### Outcome

COVID-19 vaccination status was measured starting on 23 December 2020 with two questions. First, a response (“I have already had one”) was added to our previously published^21^ study-developed item enquiring about COVID-19 vaccine intentions (“How likely do you think you are to get a COVID-19 vaccine when one is approved?”). Second, starting 8 January 2021, a second item was added: “Have you ever been offered a vaccine for COVID-19?” Response options were i) yes, twice, ii) yes, once, iii) yes, but waiting, iv) yes, but turned it down, and v) no, haven’t been offered. Our vaccination status outcome variable was constructed by classifying participants into one of two groups based on their most recent answers to these two questions: vaccinated (received at least one does or waiting) vs offered but declined.

### Potential mediators

Two variables hypothesised to mediate the association between racial/ethnic discrimination and COVID-19 vaccine refusal were considered: confidence in the central UK government and confidence in UK health service to handle the pandemic. Response options for both ranged from 1 (none at all) to 7 (lots). Two binary variables were created to compare individuals who had a lot of (5-7) versus low (1-4) confidence in the government and health system.

### Exposure

Data on experiences of racial/ethnic discrimination were collected in the last week of July 2020 with items adapted from the Everyday Discrimination Scale (EDS)^27^ which is designed to measure routine and relatively subtle experiences of unfair treatment in everyday situations. The scale is widely used and has shown expected associations with internalising and externalising symptoms.^28^ In the current study, we used seven items in total: three items from the EDS and added four questions from the English Longitudinal Study of Ageing.^29^ Participants were prompted to answer based on experiences they had had since the (first) lockdown came into effect in March 2020. We made subtle changes to some of the phrasing to account of the unique social situation of COVID-19. See Supplemental Table S2 for a full listing of item wording. Participants who said they had had each experience were asked to give one of four possible reasons (gender, race/ethnicity, age, or for another reason) for the discrimination. In the current study, the seven racial/ethnic discrimination experiences variables were summed to create a total racial/ethnic discrimination scale (0-7) with higher scores indicating more discrimination.

### Covariates

Demographic variables were measured at baseline interview: gender (male, female), education level (university degree (bachelors or higher), A-levels/equivalent or vocational, up to GCSE/O levels), and age (18-29, 30-44, 45-59, 60+). Long-term physical health condition (yes, no) using a multiple-choice question on medical conditions, also at the baseline interview. Included conditions were high blood pressure, diabetes, heart disease, lung disease, cancer, any other clinically diagnosed chronic physical health conditions, or any disability.

Having been infected with COVID-19 was categorised as a binary variable (yes, diagnosed and recovered, or yes, diagnosed and still ill, or not formally diagnosed but suspected, vs no, not that I know of or no). The presence or absence of worry about either contracting COVID-19 or becoming seriously ill from it were captured from two multiple choice questions asked during each wave of the pandemic. A binary variable was created to indicate not having endorsed either as a source of stress.

### Statistical analysis

Structural equation modelling with logistic regression was used to simultaneously test the direct and indirect effects of racial/ethnic discrimination on COVID-19 vaccine refusal through low confidence in the central UK government and in the UK health service to handle the pandemic whilst adjusting for all covariates. To increase representativeness of the UK general population, data were weighted to the proportions of gender, age, ethnicity, country, and education obtained from the UK Office for National Statistics (ONS).^30^ Analyses were conducted using Stata version 16.^31^ Coefficients were exponentiated and presented as odds ratios with 95% confidence intervals.

Sensitivity analyses were conducted with the total number of age (range 0-7), gender (range 0-7), and other (range 0-7) discrimination experiences and discrimination in medical and service settings (due to gender, race/ethnicity, age, or another reason, range 0-5) as exposures.

### Data availability

The study protocol and user guide (which includes full details on recruitment, retention, data cleaning, and sample demographics) are available at https://osf.io/jm8ra/.

## Results

The most common ethnic minority group in participants who had accepted the vaccine was Asian/Asian British (29.6%), but 13.9% of the vaccine refusal group was still comprised of this ethnic group (Table 1). Those who had declined the vaccine were over twice as likely (63.0% vs 23.9%) as those who had accepted to have a low level of education (up to GCSE/O levels) and more likely to express low confidence in the central UK government (78.9% vs 63.6%) and in the UK health service (63.8% vs 23.9%) to handle the pandemic.

**Table 1.** Descriptive characteristics of the sample by COVID-19 vaccination status (N = 633), weighted

| Variable | Total sample | At least one dose or<br>accepted/ waiting<br>% | Declined |
| --- | --- | --- | --- |
| Total sample | - | 95.92 | 4.08 |
| Ethnicity |  |  |  |
| Asian/Asian British - Indian, Pakistani, Bangladeshi, other | 28.98 | 29.62 | 13.85 |
| Black/Black British - Caribbean, Africa | 15.17 | 15.03 | 18.36 |
| Mixed race - White and Black/Black British | 9.42 | 9.76 | 1.40 |
| Mixed race - other | 25.75 | 25.83 | 23.88 |
| Chinese/Chinese British | 3.47 | 3.56 | 1.40 |
| Middle Eastern/Middle Eastern British – Arab, Turkish, other | 3.05 | 3.11 | 1.77 |
| Other ethnic group | 14.15 | 13.08 | 39.34 |
| Gender (ref male) |  |  |  |
| Female | 52.89 | 52.57 | 60.47 |
| Education (ref university degree or higher) |  |  |  |
| A-levels/equivalent or vocational | 23.70 | 24.70 | 0.00 |
| Up to GCSE/O levels | 25.49 | 23.89 | 62.95 |
| Age (ref 60+) |  |  |  |
| 45-59 | 31.26 | 31.47 | 26.36 |
| 30-44 | 23.87 | 24.22 | 15.68 |
| 18-29 | 9.11 | 9.08 | 9.87 |
| Long-term physical health condition (ref do not have) |  |  |  |
| Have a long-term physical health condition | 47.15 | 47.46 | 39.74 |
| COVID-19 illness (ref have not been infected) |  |  |  |
| Have been infected w/COVID-19 | 1.71 | 1.79 | 0.00 |
| Catching or becoming seriously ill from COVID-19 a source of stress (ref is a source of stress) |  |  |  |
| Not a source of stress | 67.42 | 67.04 | 76.24 |
| Confidence in the central UK government to handle the pandemic (ref high) |  |  |  |
| Low | 64.21 | 63.58 | 78.86 |
| Confidence in the UK health service to handle the pandemic (ref high) |  |  |  |
| Low | 25.48 | 23.85 | 63.81 |
| Racial/ethnic discrimination experiences |  |  |  |
| You have been treated with less courtesy or respect than other people | 11.02 | 10.48 | 23.67 |
| You have received poorer service than other people (e.g., for deliveries or in stores). | 3.43 | 3.45 | 3.18 |
| People have acted as if they were afraid of you | 8.16 | 7.78 | 16.95 |
| People have acted as if they think you are dishonest | 5.87 | 5.62 | 11.65 |
| You have been threatened or harassed | 2.94 | 2.91 | 3.55 |
| You have received poorer service or treatment than other people from doctors or hospitals | 1.22 | 0.98 | 6.69 |
| You have experienced some other kind of discrimination | 7.38 | 7.41 | 6.74 |
| Racial/ethnic discrimination total, M (SD) | 0.40 (1.05) | 0.39 (1.03) | 0.72 (1.41) |
| Data were weighted to the proportions of gender, age, ethnicity, country, and education obtained from the Office for National Statistics. |  |  |  |

Those who had refused the vaccine reported having experienced twice as much racial/ethnic discrimination since the start of the pandemic (M = 0.7, SD = 1.4) as those who had accepted the vaccine (M = 0.4, SD = 1.0). Nearly one in four (23.7%) in the vaccine refusal group said they had been treated with less courtesy or respect than other people because of their race/ethnicity, whilst 10.5% who had accepted the vaccine said they had. The proportion having experienced racial/ethnic discrimination in a medical setting was nearly seven times higher in the vaccine refusal than in the vaccine acceptance group (6.7% vs 0.98%).

Results from the structural equation model adjusting for covariates indicated a direct effect of racial/ethnic discrimination on low confidence in the health system to handle the pandemic (OR = 1.6, 95% CI = 1.0 to 2.0), which in turn predicted vaccine refusal (OR = 7.5, 95% CI = 2.1 to 27.4) (Table 2). There was a significant indirect effect of racial/ethnic discrimination on COVID-19 vaccine refusal via low trust in the health system (OR = 2.5, 95% CI = 1.1 to 5.4), but not low trust in government to handle the pandemic (OR = 1.0, 95% CI = 0.6 to 1.7) (Table 3). The total effect (direct and indirect via the two mediators) of racial/ethnic discrimination on COVID-19 vaccine refusal was 3.9 (95% CI = 1.4 to 10.9).

**Table 2.** Direct effects of racial/ethnic discrimination, confidence in government and the health system to handle the pandemic, and COVID-19 vaccine refusal from the structural equation model (N = 633)

|  | Low confidence in the central UK government |  |  | Low confidence in the UK health system |  |  | COVID-19 vaccine refusal |  |  |
| --- | --- | --- | --- | --- | --- | --- | --- | --- | --- |
|  | OR | 95% CI | 95% CI | OR | 95% CI | 95% CI | OR | 95% CI | 95% CI |
| Low confidence in the central UK government (ref high) | - | - | - | - | - | - | 0.94 | 0.16 | 5.45 |
| Low confidence in the UK health service (ref high) | - | - | - | - | - | - | **7.51** | **2.05** | **27.43** |
| Racial/ethnic discrimination | 1.35 | 0.94 | 1.93 | **1.56** | **1.20** | **2.04** | 1.62 | 0.96 | 2.73 |
| Model fit |  |  |  |  |  |  |  |  |  |
| Akaike's information criteria (AIC) |  |  |  |  | 2701.58 |  |  |  |  |
| Schwarz's Bayesian information criteria (BIC) |  |  |  |  | 2857.35 |  |  |  |  |
| Log pseudolikelihood |  |  |  |  | -1315.79 |  |  |  |  |

Note. Bold indicates $p < 0.05$ . Data were weighted to the proportions of gender, age, ethnicity, country, and education obtained from the Office for National Statistics. Structural equation model adjusts for COVID-19 infection status, not being worried about catching or becoming seriously ill from COVID-19, gender, age, education level, and the presence of a long-term physical health condition.

**Table 3.** Indirect effects of racial/ethnic discrimination, confidence in government and the health system to handle the pandemic, and COVID-19 vaccine refusal from the structural equation model (N = 633)

|  | COVID-19 vaccine refusal |  |  |
| --- | --- | --- | --- |
|  | OR | 95% CI | 95% CI |
| Indirect effect of racial/ethnic discrimination on COVID-19 vaccine refusal through low confidence in the central UK government to handle the pandemic | 0.98 | 0.58 | 1.67 |
| Indirect effect of racial/ethnic discrimination on COVID-19 vaccine refusal through low confidence in the UK health service to handle the pandemic | **2.46** | **1.12** | **5.39** |
| Total effect of racial/ethnic discrimination on COVID-19 vaccine refusal (direct effect of racial/ethnic discrimination plus indirect effects of low confidence in government and the health system) | **3.91** | **1.40** | **10.92** |

Note. Bold indicates $p < 0.05$ . Data were weighted to the proportions of gender, age, ethnicity, country, and education obtained from the Office for National Statistics.

Sensitivity analyses indicated indirect effects of age (Tables S3 and S4) and gender discrimination (Tables S5 and S6) on COVID-19 vaccine refusal through low trust in the health system. Neither discrimination taking place in medical nor service settings (due to gender, race, age, or other) had direct or indirect effects on vaccine refusal.

## Discussion

This is the first study in the UK, to our knowledge, that finds longitudinal associations between racial/ethnic discrimination and COVID-19 vaccine refusal. The total effect of racial/ethnic discrimination on vaccine refusal was nearly four-fold. This echoes previous research showing associations between lifetime experiences of racial discrimination with COVID-19 vaccine hesitancy.^14^ Our study expands on previous research by showing that low trust in the health system mediates the relationship between racial/ethnic discrimination and vaccine refusal. Further, our findings confirm evidence before the current pandemic, which found associations between experiences of racial discrimination and distrust of the health care system^32,33^ and physicians^34^ among ethnic minority adults. Recent research also finds widespread racial and ethnic discrimination towards ethnic minority healthcare professionals within the UK National Health Service (NHS).^35^ Together, these findings highlight the crucial role of the institutions, in particular the NHS, in building and maintaining trust in the affected communities.

In this study, 6.7% of participants who had refused the vaccine reported they had experienced poorer service or treatment than other people in a medical setting because of their race or ethnicity. Research conducted before and during the pandemic also suggests that lifetime experiences of racial/ethnic discrimination in health care settings is commonplace. In June 2019, nearly 1 in 3 (30%) of non-Hispanic Black and over 1 in 10 (11%) Hispanic adults had ever been treated differently by a health care provider because of their race or ethnicity.^18^ Mistreatment by a doctor or nurse due to race was also reported by nearly one in ten (9%) of respondents in a US study conducted at the end of December 2020.^14^ Small studies of ethnic minority adults suggest that not feeling listened to by medical professionals may be a particularly common experience of discrimination in medical settings.^33,36,37^ Future studies should seek to identify specific situations and settings in which this type of discrimination is most likely to take place in the context of the COVID-19 pandemic.

We examined the total number of racial/ethnic discrimination experiences in relation to vaccine refusal and it was low trust in the health system, and not the central UK government, to handle the pandemic that mediated the association between discrimination and vaccine refusal. Research, in mostly White participants, has found similar associations between low trust in the health system, not central government, and negative attitudes towards vaccines, in particular COVID-19 vaccination.^21^ Therefore, building trust in the healthcare system will be key for effective management of the current and future pandemics as well as public health campaigns in general.

Based on our findings, focusing exclusively on vaccine misinformation may disregard concerns about mistrust that is largely due to past experiences of racial and ethnic discrimination. Public health messaging that communicate vaccine safety should also be delivered in multiple languages by trusted and relatable sources of information to increase their reach and effectiveness.^20^

## Strengths and limitations

Strengths of this study include a longitudinal design and a large sample size that includes a diverse group of UK ethnic minority adults from different age, gender, socioeconomic status, and geographical locations. Although data were weighted to increase representativeness of the general UK population, sampling was not random, and caution should therefore be used in generalising results. Notably, the proportion of ethnic minority adults in our study who had refused the COVID-19 vaccine was less than half that reported by the ONS (4.1% vs 9%).^1^ Due to the small number of participants within specific ethnic minority groups in our sample, we examined ethnic minority groups as a whole in our analyses, which does not account for the variation in the current and historical experiences of discrimination, as well as any underlying differences in reasons for vaccine hesitancy due to age, sex, and education across these diverse groups.^3,20^ Further, due to limitations in question phrasing (i.e.,‘White-British, Irish, other’), we were unable to examine associations between study variables and vaccine refusal in White subgroups, some of whom have also had lower COVID-19 vaccine uptake.^38^

The most common ethnic group in those who had refused the vaccine was the ‘other ethnic group’. Similarly, nearly one in ten (7.4%) in the total sample said they had experienced some ‘other’ form of discrimination related to their race or ethnicity. Future research should therefore provide participants with opportunities to write in their identified ethnic group and specify other types of racial discrimination experienced. Furthermore, future studies should collect information on the frequency of racial/ethnic discrimination to gain understanding of the extent and magnitude of the experiences of daily discrimination. Finally, another limitation of the current study is that we measured trust in government and the health service, and not mistrust, the latter of which implies beliefs that institutions are actively behaving in contradiction of one’s best interests.^19^

## Conclusion

The adverse effects of racial/ethnic discrimination on health and health outcomes in marginalised ethnic groups are well-established in the literature.^13,39^ Our study builds upon existing evidence that racial discrimination increases COVID-19 vaccine hesitancy ^14^ by demonstrating that a nearly four-fold effect of racial discrimination on vaccine refusal is mediated by low trust in the health system. These findings indicate that it is vital that healthcare institutions such as the NHS work to gain the confidence and trust of ethnic minority groups. Furthermore, public health campaigns to increase COVID-19 uptake in ethnic minorities should include not only trust-building in vaccines, but also strategies to prevent racial discrimination and support ethnic minorities who have experienced discrimination.

## Data Availability

The study protocol and user guide (which includes full details on recruitment, retention, data cleaning, and sample demographics) are available at https://github.com/UCL-BSH/CSSUserGuide.

## Acknowledgements

We would like to extend our gratitude to participants in the UCL COVID-19 Social Study as well as other researchers working on the study.

## Supplemental Materials

**Table S1.**
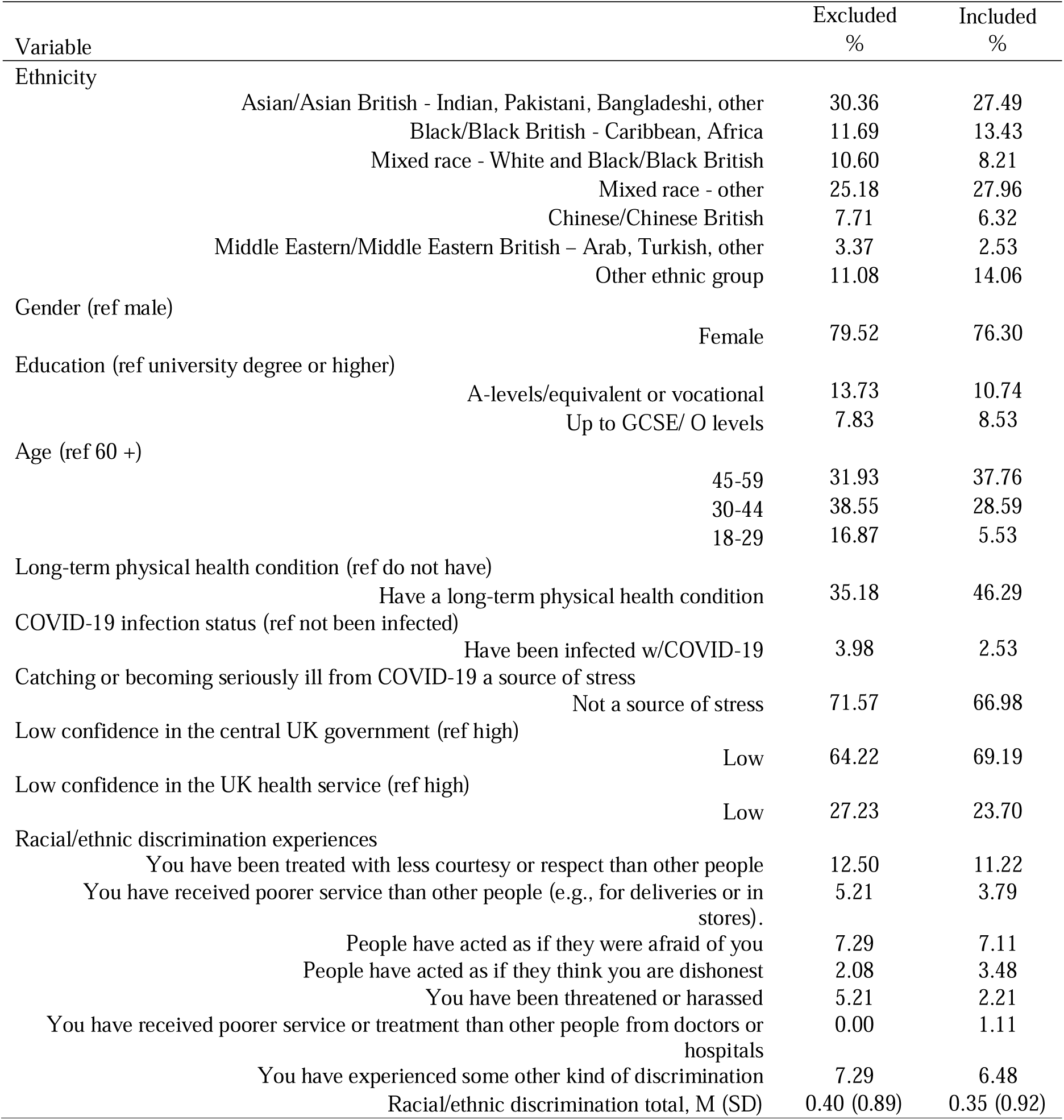
Characteristics of included and excluded participants, unweighted

**Table S2.**
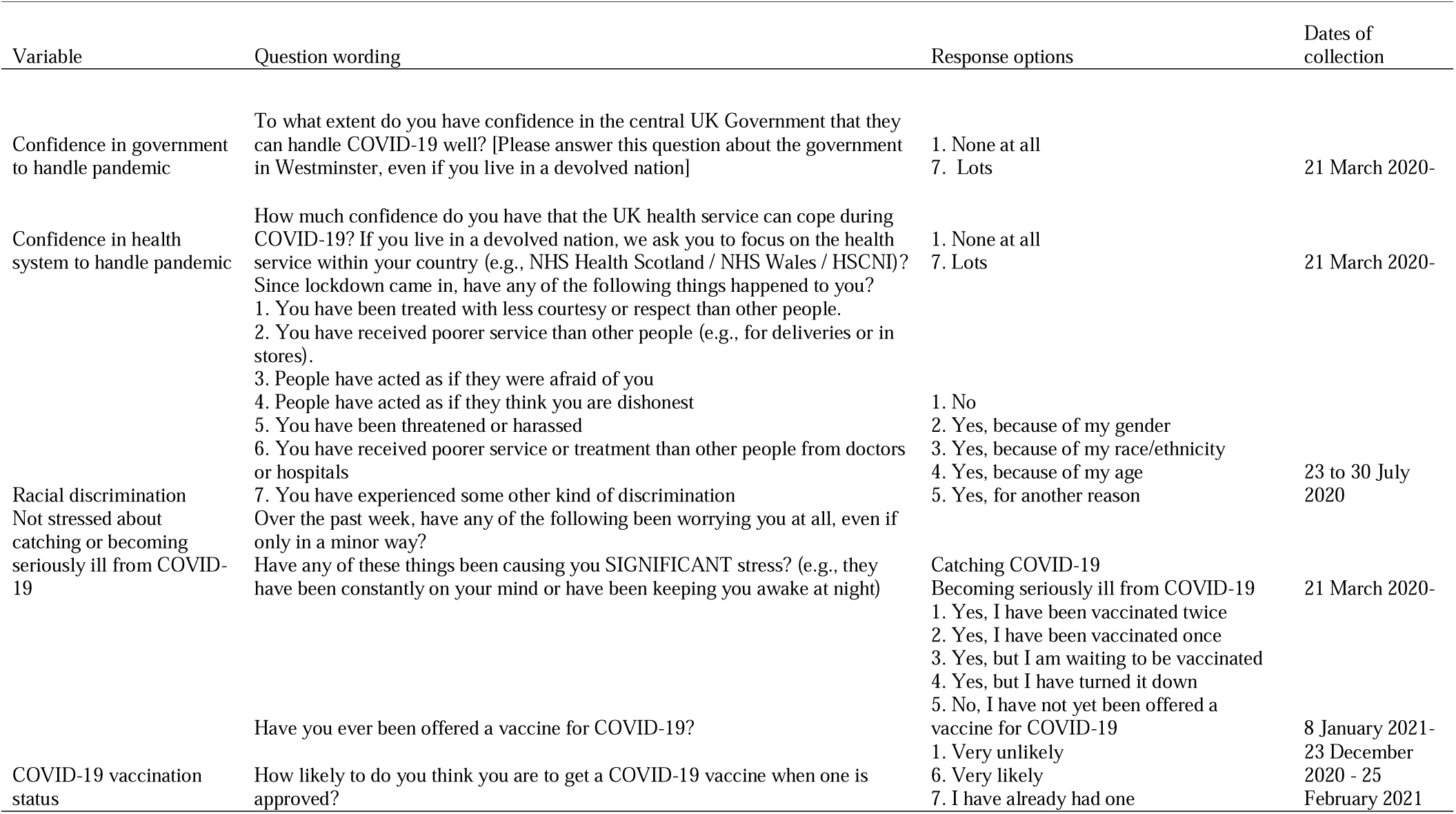
Wording of study-developed and modified items

**Table S3.**
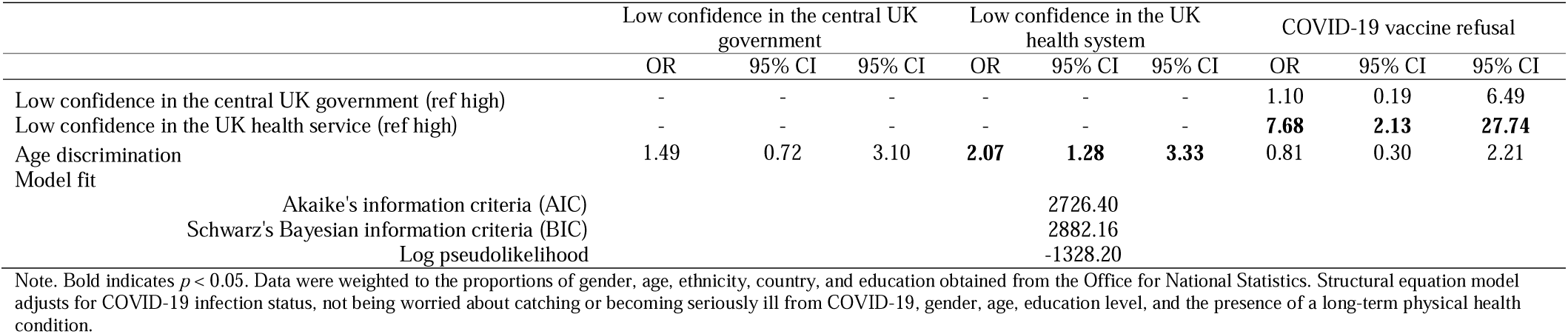
Sensitivity analysis: Direct effects of age discrimination, confidence in government and the health system and COVID-19 vaccine refusal from the structural equation model (N = 633)

**Table S4.** Sensitivity analysis: Indirect effects of age discrimination, confidence in government and the health system and COVID-19 vaccine refusal from the structural equation model (N = 633)

|  | COVID-19 vaccine refusal |  |  |
| --- | --- | --- | --- |
|  | OR | 95% CI | 95% CI |
| Indirect effect of age discrimination on COVID-19 vaccine refusal through low confidence in the central UK government | 1.04 | 0.51 | 2.11 |
| Indirect effect of age discrimination on COVID-19 vaccine refusal through low confidence in the UK health service | **4.41** | **1.16** | **16.74** |
| Total effect of age discrimination on COVID-19 vaccine refusal (direct effect of age discrimination plus indirect effects of low confidence in government and the health system) | 3.73 | 0.71 | 19.65 |

Note. Bold indicates $p < 0.05$ . Data were weighted to the proportions of gender, age, ethnicity, country, and education obtained from the Office for National Statistics. Structural equation model adjusts for COVID-19 infection status, not being worried about catching or becoming seriously ill from COVID-19, gender, age, education level, and the presence of a long-term physical health condition.

**Table S5.** Sensitivity analysis: Direct effects of gender discrimination, confidence in government and the health system and COVID-19 vaccine refusal from the structural equation model (N = 633)

|  | Low confidence in the central UK government |  |  | Low confidence in the UK health system |  |  | COVID-19 vaccine refusal |  |  |
| --- | --- | --- | --- | --- | --- | --- | --- | --- | --- |
|  | OR | 95% CI | 95% CI | OR | 95% CI | 95% CI | OR | 95% CI | 95% CI |
| Low confidence in the central UK government (ref high) | - | - | - | - | - | - | 1.07 | 0.18 | 6.42 |
| Low confidence in the UK health service (ref high) | - | - | - | - | - | - | **6.89** | **1.96** | **24.16** |
| Gender discrimination | 1.85 | 0.83 | 4.15 | **2.84** | **1.43** | **5.65** | 1.61 | 0.90 | 2.89 |
| Model fit |  |  |  |  |  |  |  |  |  |
| Akaike's information criteria (AIC) |  |  |  |  |  | 2715.932 |  |  |  |
| Schwarz's Bayesian information criteria (BIC) |  |  |  |  |  | 2871.699 |  |  |  |
| Log pseudolikelihood |  |  |  |  |  | -1322.97 |  |  |  |

Note. Bold indicates $p < 0.05$ . Data were weighted to the proportions of gender, age, ethnicity, country, and education obtained from the Office for National Statistics. Structural equation model adjusts for COVID-19 infection status, not being worried about catching or becoming seriously ill from COVID-19, gender, age, education level, and the presence of a long-term physical health condition.

**Table S6.** Sensitivity analysis: Indirect effects of gender discrimination, confidence in government and the health system and COVID-19 vaccine refusal from the structural equation model (N = 633)

|  | COVID-19 vaccine refusal |  |  |
| --- | --- | --- | --- |
|  | OR | 95% CI | 95% CI |
| Indirect effect of gender discrimination on COVID-19 vaccine refusal through low confidence in the central UK government | 1.04 | 0.35 | 2.15 |
| Indirect effect of gender discrimination on COVID-19 vaccine refusal through low confidence in the UK health service | **7.49** | **1.11** | **50.50** |
| Total effect of gender discrimination on COVID-19 vaccine refusal (direct effect of gender discrimination plus indirect effects of low confidence in government and the health system) | **12.57** | **1.37** | **115.58** |

Note. Bold indicates $p < 0.05$ . Data were weighted to the proportions of gender, age, ethnicity, country, and education obtained from the Office for National Statistics. Structural equation model adjusts for COVID-19 infection status, not being worried about catching or becoming seriously ill from COVID-19, gender, age, education level, and the presence of a long-term physical health condition.

**Table S7.** Sensitivity analysis: Direct effects of other discrimination, confidence in government and the health system and COVID-19 vaccine refusal from the structural equation model (N = 633)

|  | Low confidence in the central UK government |  |  | Low confidence in the UK health system |  |  | COVID-19 vaccine refusal |  |  |
| --- | --- | --- | --- | --- | --- | --- | --- | --- | --- |
|  | OR | 95% CI | 95% CI | OR | 95% CI | 95% CI | OR | 95% CI | 95% CI |
| Low confidence in the central UK government (ref high) | - | - | - | - | - | - | 1.08 | 0.18 | 6.69 |
| Low confidence in the UK health service (ref high) | - | - | - | - | - | - | **6.76** | **1.89** | **24.18** |
| Other discrimination | 1.27 | 0.98 | 1.64 | 1.18 | 0.87 | 1.60 | **1.59** | **1.11** | **2.26** |
| Model fit |  |  |  |  |  |  |  |  |  |
| Akaike's information criteria (AIC) |  |  |  |  |  |  | 2737.81 |  |  |
| Schwarz's Bayesian information criteria (BIC) |  |  |  |  |  |  | 2893.57 |  |  |
| Log pseudolikelihood |  |  |  |  |  |  | -1333.90 |  |  |

Note. Bold indicates $p < 0.05$ . Data were weighted to the proportions of gender, age, ethnicity, country, and education obtained from the Office for National Statistics. Structural equation model adjusts for COVID-19 infection status, not being worried about catching or becoming seriously ill from COVID-19, gender, age, education level, and the presence of a long-term physical health condition.

**Table S8.** Sensitivity analysis: Indirect effects of other discrimination, confidence in government and the health system and COVID-19 vaccine refusal from the structural equation model (N = 633)

|  | COVID-19 vaccine refusal |  |  |
| --- | --- | --- | --- |
|  | OR | 95% CI | 95% CI |
| Indirect effect of other discrimination on COVID-19 vaccine refusal through low confidence in the central UK government | 1.01 | 0.75 | 1.36 |
| Indirect effect of other discrimination on COVID-19 vaccine refusal through low confidence in the UK health service | 1.58 | 0.86 | 2.90 |
| Total effect of other discrimination on COVID-19 vaccine refusal (direct effect of other discrimination plus indirect effects of low confidence in government and the health system) | **2.54** | **1.09** | **5.89** |

Note. Bold indicates $p < 0.05$ . Data were weighted to the proportions of gender, age, ethnicity, country, and education obtained from the Office for National Statistics. Structural equation model adjusts for COVID-19 infection status, not being worried about catching or becoming seriously ill from COVID-19, gender, age, education level, and the presence of a long-term physical health condition.

**Table S9.** Sensitivity analysis: Direct effects of discrimination in medical settings, confidence in government and the health system and COVID-19 vaccine refusal from the structural equation model (N = 633)

|  | Low confidence in the central UK government |  |  | Low confidence in the UK health system |  |  | COVID-19 vaccine refusal |  |  |
| --- | --- | --- | --- | --- | --- | --- | --- | --- | --- |
|  | OR | 95% CI | 95% CI | OR | 95% CI | 95% CI | OR | 95% CI | 95% CI |
| Low confidence in the central UK government (ref high) | - | - | - | - | - | - | 1.11 | 0.19 | 6.57 |
| Low confidence in the UK health service (ref high) | - | - | - | - | - | - | **8.06** | **2.22** | **29.22** |
| Discrimination in medical settings | 0.55 | 0.21 | 1.44 | 0.91 | 0.33 | 2.45 | 3.05 | 0.90 | 10.27 |
| Model fit |  |  |  |  |  |  |  |  |  |
| Akaike's information criteria (AIC) |  |  |  |  |  |  | 2748.78 |  |  |
| Schwarz's Bayesian information criteria (BIC) |  |  |  |  |  |  | 2904.55 |  |  |
| Log pseudolikelihood |  |  |  |  |  |  | -1339.39 |  |  |

Note. Bold indicates $p < 0.05$ . Data were weighted to the proportions of gender, age, ethnicity, country, and education obtained from the Office for National Statistics. Structural equation model adjusts for COVID-19 infection status, not being worried about catching or becoming seriously ill from COVID-19, gender, age, education level, and the presence of a long-term physical health condition.

**Table S10.** Sensitivity analysis: Indirect effects of discrimination in medical settings, confidence in government and the health system and COVID-19 vaccine refusal from the structural equation model (N = 633)

|  | COVID-19 vaccine refusal |  |  |
| --- | --- | --- | --- |
|  | OR | 95% CI | 95% CI |
| Indirect effect of discrimination in medical settings on COVID-19 vaccine refusal through low confidence in the central UK government | 0.94 | 0.32 | 2.77 |
| Indirect effect of discrimination in medical settings on COVID-19 vaccine refusal through low confidence in the UK health service | 0.81 | 0.10 | 6.54 |
| Total effect of discrimination in medical settings on COVID-19 vaccine refusal (direct effect of discrimination in medical settings plus indirect effects of low confidence in government and the health system) | 2.33 | 0.15 | 35.27 |

Note. Bold indicates $p < 0.05$ . Data were weighted to the proportions of gender, age, ethnicity, country, and education obtained from the Office for National Statistics. Structural equation model adjusts for COVID-19 infection status, not being worried about catching or becoming seriously ill from COVID-19, gender, age, education level, and the presence of a long-term physical health condition.

**Table S11.** Sensitivity analysis: Direct effects of discrimination in service settings, confidence in government and the health system and COVID-19 vaccine refusal from the structural equation model (N = 633)

|  | Low confidence in the central UK government |  |  | Low confidence in the UK health system |  |  | COVID-19 vaccine refusal |  |  |
| --- | --- | --- | --- | --- | --- | --- | --- | --- | --- |
|  | OR | 95% CI | 95% CI | OR | 95% CI | 95% CI | OR | 95% CI | 95% CI |
| Low confidence in the central UK government (ref high) | - | - | - | - | - | - | 1.11 | 0.19 | 6.57 |
| Low confidence in the UK health service (ref high) | - | - | - | - | - | - | **8.06** | **2.22** | **29.22** |
| Discrimination in service settings | 0.91 | 0.33 | 2.45 | 0.55 | 0.21 | 1.44 | 3.05 | 0.90 | 10.27 |
| Model fit |  |  |  |  |  |  |  |  |  |
| Akaike's information criteria (AIC) |  |  |  |  | 2704.42 |  |  |  |  |
| Schwarz's Bayesian information criteria (BIC) |  |  |  |  | 2860.19 |  |  |  |  |
| Log pseudolikelihood |  |  |  |  | -1317.21 |  |  |  |  |

Note. Bold indicates $p < 0.05$ . Data were weighted to the proportions of gender, age, ethnicity, country, and education obtained from the Office for National Statistics. Structural equation model adjusts for COVID-19 infection status, not being worried about catching or becoming seriously ill from COVID-19, gender, age, education level, and the presence of a long-term physical health condition.

**Table S12.** Sensitivity analysis: Indirect effects of discrimination in service settings, confidence in government and the health system and COVID-19 vaccine refusal from the structural equation model (N = 633)

|  |
| --- |
| Indirect effect of discrimination in service settings on COVID-19 vaccine refusal through low confidence in the central UK governr |
| Indirect effect of discrimination in service settings on COVID-19 vaccine refusal through low confidence in the UK health service |
| Total effect of discrimination in service settings on COVID-19 vaccine refusal (direct effect of discrimination in service settings plus indirect effects of low confidence in government and the health system) |

Note. Bold indicates $p < 0.05$ . Data were weighted to the proportions of gender, age, ethnicity, country, and education obtained from the Office for National Statistics. Structural equation model adjusts for COVID-19 infection status, not being worried about catching or becoming seriously ill from COVID-19, gender, age, education level, and the presence of a long-term physical health condition.

